# Comparison of external treatment of Acupuncture and moxibustion and intervention of Chinese and Western Medicine on postoperative pain of hemorrhoids: A systematic review and meta-analysis

**DOI:** 10.1101/2023.01.20.23284830

**Authors:** XinYan Zou, QiaoQiao Liu, LongXia Gao, HanQing Zhao

## Abstract

**Objective:** To evaluate the clinical efficacy and safety of acupuncture and moxibustion in the treatment of postoperative pain of hemorrhoids compared with traditional Chinese medicine and western medicine.

**Methods:** The CNKI, pubMed, Cochrane Library, Science Direct, Wan Fang, VIP, CBM, WOS, Bailian Yun Library and other databases were systematically retrieved from 2017 to October 2022 for clinical randomized controlled trials of acupuncture versus traditional Chinese and Western medicine for postoperative pain in hemorrhoids. The two evaluators independently retrieved, sifted through literature and extracted data for inclusion in a randomized controlled trial of acupuncture for the treatment of hemorrhoid pain that matched the study. Literature quality assessment was performed using RevMan5.4 for meta-analysis.

**Results:** A total of 540 related literature articles were retrieved, of which 139 were from CNKI, 104 from Wan Fang, 104 from VIP26, 7 from PubMed, 9 from Cochrane, 35 from WOS, 173 from China Biomedical Literature Database, 1 from Science Direct and 46 from the Bailian Yun Library, Screening resulted in inclusion of 10 RCTs including 870 patients. Meta analysis showed that there was no significant difference in the degree of pain in 2 hours [MD=0.01, 95%CI (−0.23, 0.24), P ≤ 0.95]. And it showed that the total effective rate of the two groups was [RR=1.14, 95%CI (1.06, 1.24), P ≤ 0.0001], intervention for 2days pain degree was [MD=-0.41, 95%CI (−0.69, 0.13), P ≤ 0.004], the incidence of adverse reaction was [RR=0.15, 95%CI (0.03, 0.79), P=0.03], the difference was statistically significant (P<0.05).

**Conclusion:** Drug treatment is effective quickly, analgesia effect is better than acupuncture in early treatment, but the effect is not lasting. Acupuncture treatment is slow to start but the effects of acupuncture will gradually become apparent at a later stage. However, due to the low quality of inclusion, multicenter, large sample size and double-blind randomized controlled trials are still needed.

---

Hemorrhoids are soft clusters of veins or connective tissue, commonly known as hemorrhoids, that form when the veins at the end of the rectum become convoluted and expand under the skin of the anus, or when blood clots or growths form under the skin of the anus. According to TCM, the disease is caused by deficiency of viscera, sedentary and heavy travel. Or eating disorders, eating spicy, fat; or chronic constipation, diarrhea; Or tired, fetal birth and so on can lead to anal qi and blood imbalance, stagnant complexes, contains ShiRe then cause dysentery. Hemorrhoid is the common, multiple diseases, therefore the folk have “ten people nine hemorrhoids’’. Western medicine says a low-fiber diet[1] and constipation are also important causes of hemorrhoids. Studies have also shown[2] that sugar can reduce hemorrhoid edema and inflammation. Surgery[3][4] was the main treatment[5][6], but there were more complications such as pain[7], edema, constipation and itching, which affected the quality of life and recovery. This paper focuses on postoperative pain as a complication of systematic discussion. There are two kinds of causes of pain according to traditional chinese medicine: One is unobstructed, the other is weakness. Here is the hemorrhoid surgery caused by incision injury, damage to the tendons and collaterals, qi and blood stasis, the meridians are not smooth, then pain. Treatment is suitable for clearing heat and detoxification, activating blood circulation and removing blood stasis, reducing swelling and pain. Acupuncture has long been used to treat hemorrhoids. The experience of acupuncture in treating hemorrhoids and acupoints have been recorded in Huangdi Neijing (The Yellow Emperor’s Inner Classic). Jin Dynasty “Zhenjiu Jiayijing” said:”Hemorrhoids pain, treat with cuanzhu; hemorrhoids, treat with huiyin.” Acupuncture and moxibustion[8][9] has unique advantages in relieving pain after hemorrhoid surgery. It has the advantages of unclogging meridians and collaterals, invigorating blood flow and relieving pain. At present, the clinical research on acupuncture and moxibustion for postoperative pain of hemorrhoids is obviously insufficient, there is a small sample size and no uniform evaluation index, so this study uses meta analysis to evaluate the effect of acupuncture on postoperative pain complications of hemorrhoids compared with traditional chinese medicine[10] and western medicine. In order to provide a reference for the clinical study of acupuncture and moxibustion in the treatment of postoperative complications of anal intestinal diseases.

## 1. Information and methodology

### 1.1 Inclusion and exclusion criteria

1.1.1 Inclusion criteria ① study type: randomized controlled trial (RCT); ② Subjects: postoperative patients who met the criteria for diagnosis of mixed hemorrhoid disease, excluding children, pregnant and lactating women; ③ Intervention: the observation group uses acupuncture or moxibustion treatment, the specific position or the time limit is not limited; The control group was treated with western medicine or traditional Chinese medicine, and the specific medicine or dosage form was unlimited. ④ Outcome indicators: total efficacy, degree of pain, incidence of adverse reactions.

1.1.2 Exclusion criteria ①Non-Chinese and English literature; ②Non-randomized controlled trials; ③ Literature in association with traditional Chinese medicine; ④ Republished literature; ⑤ Documents for which raw data are unavailable or incomplete and for which quality is low.

### 1.2 Document retrieval strategy

Computerized retrieval of CNKI, pubMed, Cochrane Library, Science Direct, WanFang, VIP, CBM, WOS, Bailian Yun Library and others from 2017 to October 2022 for clinical randomized controlled trials of acupuncture versus traditional Chinese and Western medicine for postoperative pain in hemorrhoids. No related unpublished academic papers and conference papers were collected. In order to retrieve it more comprehensively, chinese search words using “痔疮” “痔病” “痔核” “混合痔” “内痔” “外痔” “疼痛” “针灸” “刺灸疗法” “针疗法” “针灸疗法” “电”, English search words using “hemorrhoide” “hemorrhoides” “splitting pains” “crushing pains” “ache” “pain” “pharmacopuncture” “acupuncture”. And according to the characteristics of each database, subject words and free words are used for comprehensive retrieval. Take CNKI for example, the specific retrieval strategy is(: 痔疮 + 痔核 + 痔病 + 内痔 + 混合痔) AND(针灸 + 刺灸疗法 + 针疗法 + 针灸疗法 + 电针) AND 疼痛?

### 1.3 Documentation screening and information extraction

The titles and abstracts of the required literature were obtained from a complete searchable database and introduced into the NoteExpress document management software. After reading the titles and abstracts, a preliminary screen was conducted to exclude literature that clearly did not fit the study. A full text of the literature that might fit the study was then downloaded, references to the literature were extracted and registered using Excel forms. The abstractions included the name of the researcher, the year of publication, the general characteristics of the patient and sample size, intervention measures, and efficacy indicators.

### 1.4 Bias Risk Assessment Included in the Study

This study utilizes the RCT Bias Risk Assessment Tool provided by Cochrane, which evaluates randomized sequence generation; Allocation of concealment; Implementation of blind law (patients, researchers); The implementation of blind method in outcome evaluation; Results Complete data; Selective publication of reports; Other biases. The risk of bias in the literature was assessed at three levels: low risk, high risk and uncertainty. These sections are operated independently by two researchers according to the same criteria, and if contested literature is encountered, a joint decision is made through discussion with a third researcher.

### 1.5 Statistical Analysis

Meta-analysis was performed using RevMan. Relative risk (RR) was used as an effect indicator for the dichotomous data, and a 95% CI was given for the continuous data using standardized mean difference (SMD) or weighted mean difference (WMD). Q-test and I ^2^ values were used to include study heterogeneity, if there was no statistical difference (P > 0.1, I ^2^ < 50%), fixed effect models were used for Meta analysis; if there was statistical heterogeneity between studies (P ≤ 0.1, I ^2^ ≥ 50%), further analysis of heterogeneity sources was performed, and after excluding significant clinical heterogeneity, randomized effect models were used for Meta analysis. Obvious clinical heterogeneity was treated with subgroup analysis or only for profiling analysis. Incorporating funnel graph analysis into the published bias of the study, the difference was statistically significant at P < 0.05.

## 2. Result

### 2.1 Documentation screening process and results

Initial examination yielded 106 relevant literature articles, which were eventually incorporated into 10 literature articles for quantitative synthetic meta analysis, including 870 patients. The process and results of literature screening are shown in Figure 1.

**Figure 1.**
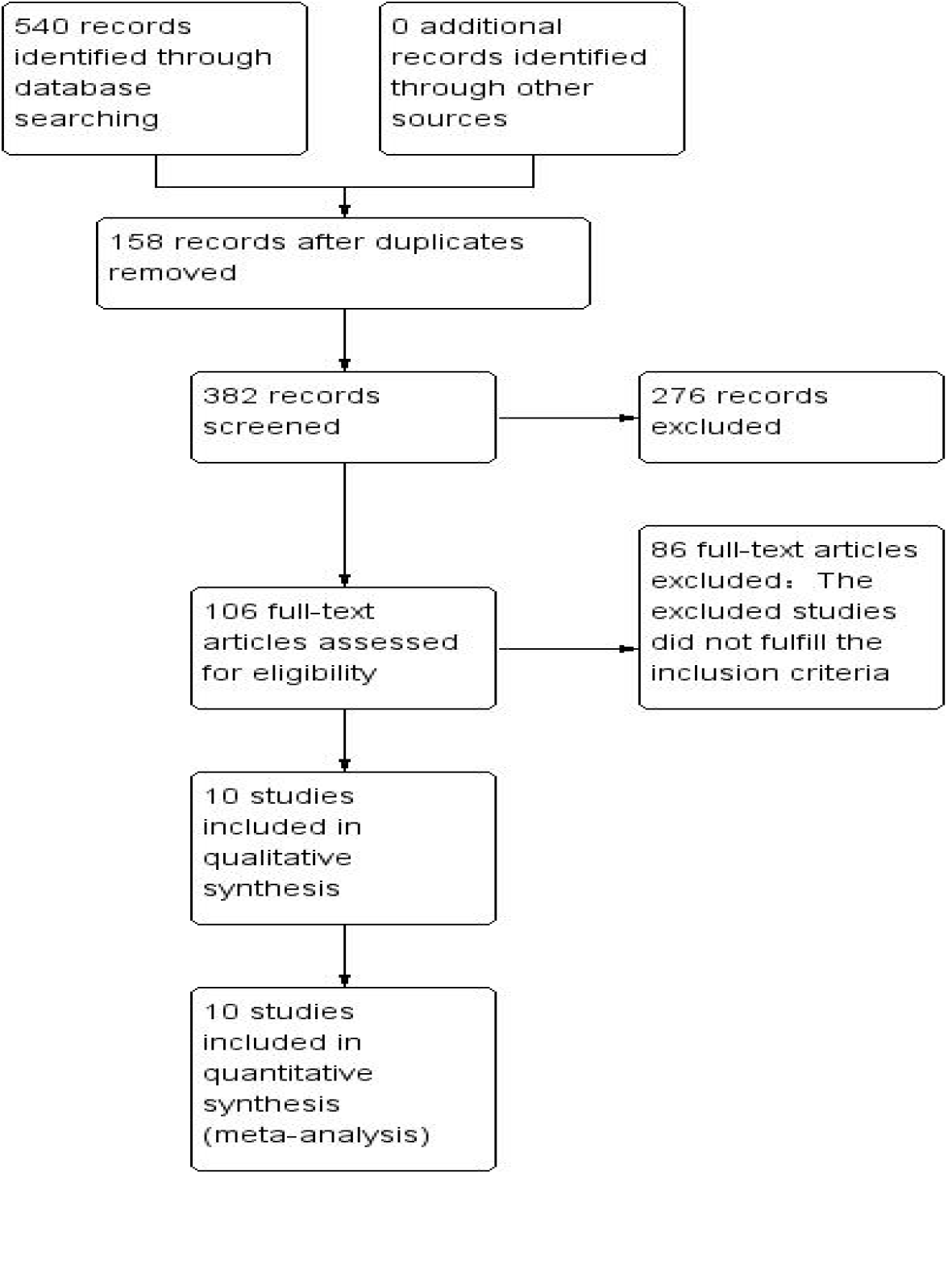
Study screening process.

### 2.2 Include basic features of the study and Quality assessment

The basic features included in the study are presented in table 1. Quality assessments are shown in figure 2.

**Table 1.** Basic features of inclusion research

| serial<br>number | First Author and Year | Type of study | Sample (T / C) | Age (T / C, years) | Interventions |  | Period of<br>treatment (d) |
| --- | --- | --- | --- | --- | --- | --- | --- |
|  |  |  |  |  | T | C |  |
| 1 | Gu Jiao 2020[11] | Random<br>Numbers Table | 49/49 | 25-55/35-65 | Acupuncture (once/<br>day,retaining 4-6h) | Analgesics, ketoclopyrate<br>ambutriol capsules (twice/day,10mg<br>each time) | 14 |
| 2 | Zu Ruihong2018[12] | Random<br>Numbers Table | 60/60 | 30-46/32-45 | Acupoint application,<br>acupuncture | Sufentanil, flubilophene,<br>toproxytrone, saline | 2 |
| 3 | He Yinghua2018[13] | Random<br>Numbers Table | 40/40 | 47.1±10/46±6.8 | Ear acupressure press | Aminophenoxycodone tablets | Not<br>mentioned |
| 4 | Sun Jiejing2020[14] | Random<br>Numbers Table | 29/30 | 18-60 | Acupuncture (once/<br>day,retaining 10min) | Aminotramadol tablets (once/ day,<br>1 tablet / session) | 3 |
| 5 | Li Kaili2021[15] | Random<br>Numbers Table | 36/36 | 18-65 | Acupuncture of<br>AnshenLiqi method | indomethacin sustained-release<br>capsules | 5 |
| 6 | Zhang Zihao2021[16] | Random<br>Numbers Table | 35/35 | 42.03±11.11/<br>43.57±11.42 | Electroacupuncture | Sodium dichlorophenol | 3 |
| 7 | Lei Hongfeng2019[17] | Random<br>Numbers Table | 30/30 | 45.9±5.8/46.4<br>±6.3 | Acupuncture and Heat<br>Moxibustion | Anal lotion with JinHuang cream | 7 |
| 8 | Dai Junmei2019[18] | Random<br>Numbers Table | 14/17 | 46.8±5.3/47.9<br>±5.2 | Acupoint application<br>and moxibustion treatment | Ketonoptyrolysis Ammontriol<br>Injection, Sodium Chloride Injection | Not<br>mentioned |

Note: T: Observation Group; C: Control group; ①: Total Efficiency②: NRS Pain Score ③: Patient Satisfaction ④: VAS Pain Score ⑤: Incidence of Adverse Effects ⑥: Quality of Life Score ⑦: Urinary retention Score ⑧: Anal Edema Score ⑨: Hospital Time

**Figure 2.**
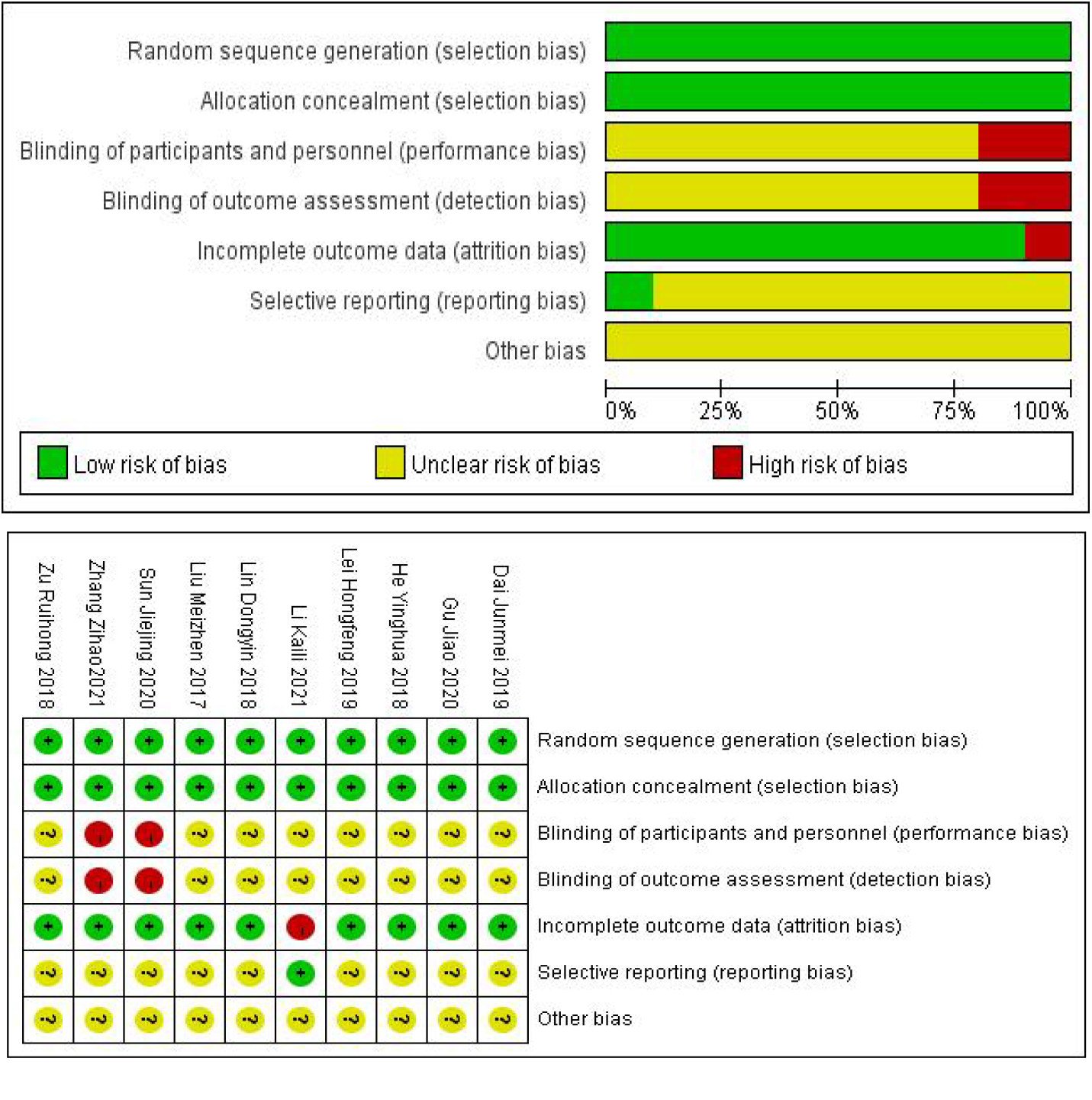
Quality Assessment

### 2.3 Meta Analysis

#### 2.3.1 overall clinical efficacy

The total efficacy of clinical efficacy was reported in six literature studies[11][13][14][15][16][19] with no significant heterogeneity between the studies (heterogeneity assays P=0.49, I^2^=0) and combined analysis using fixed-effect models showed statistically significant differences between the two groups [RR=1.14, 95% CI (1.06, 1.24), P = 0.001], as shown in Figure 3.

**Figure 3.**
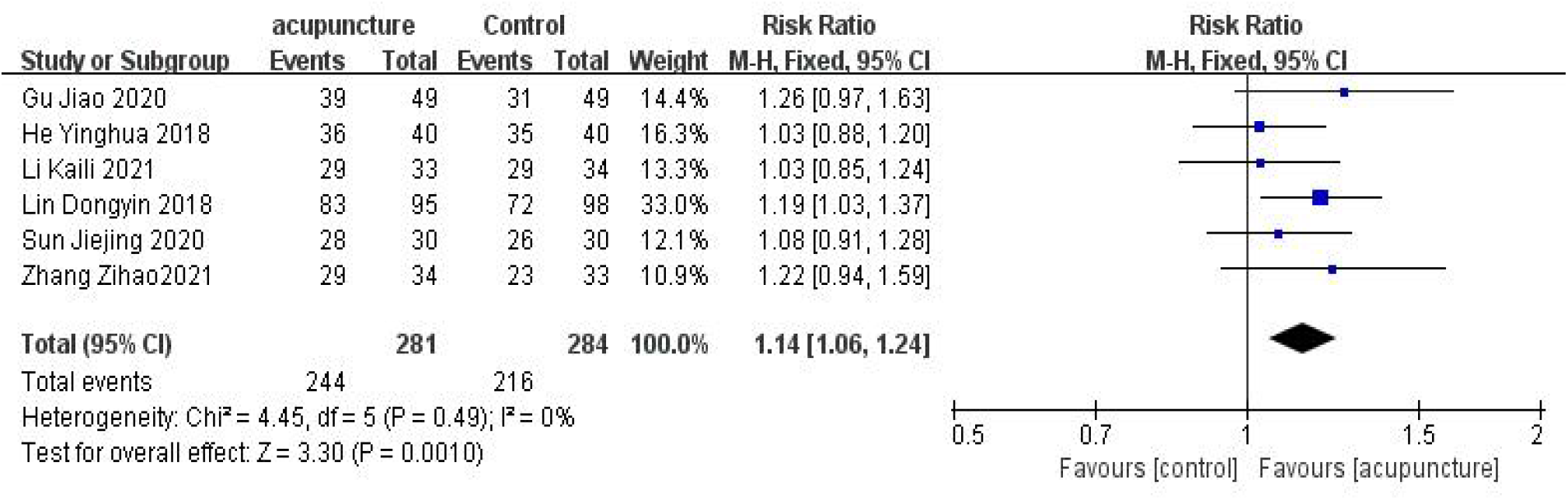
Forest plot for total Clinical Efficiency

#### 2.3.2 degree of pain

Pain levels were reported in 3 literature studies[12][16][18] 2 hours after the intervention, with no significant heterogeneity between the studies (heterogeneity assay P=0.95, I ^2^ =23%). Combined analysis using fixed-effect models showed no statistically significant difference between the two groups in pain levels 2 hours after the intervention [MD=0.01, 95% CI (−0.23, 0.24), P=0.95% CI), as shown in figure 4. Two studies[14][16]reported pain levels in patients two days after the intervention, with no significant heterogeneity between studies (heterogeneity assays P=0.004, I^2^ =0). Combined analysis using fixed-effect models showed that MD=-0.41, 95% CI (−0.69, -0.13) and two groups had statistically significant differences in pain levels after two days of intervention [MD=-0.41, 95% CI (−0.69, -0.13), P=0.004], as shown in Figure 5.

**Figure 4.**
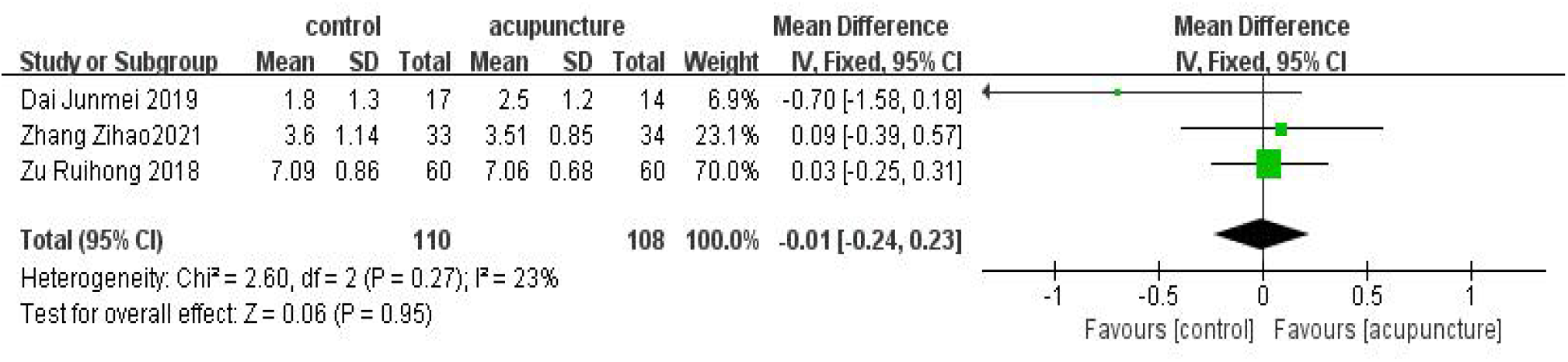
Forest plot of 2-hour postoperative intervention pain levels

**Figure 5.**
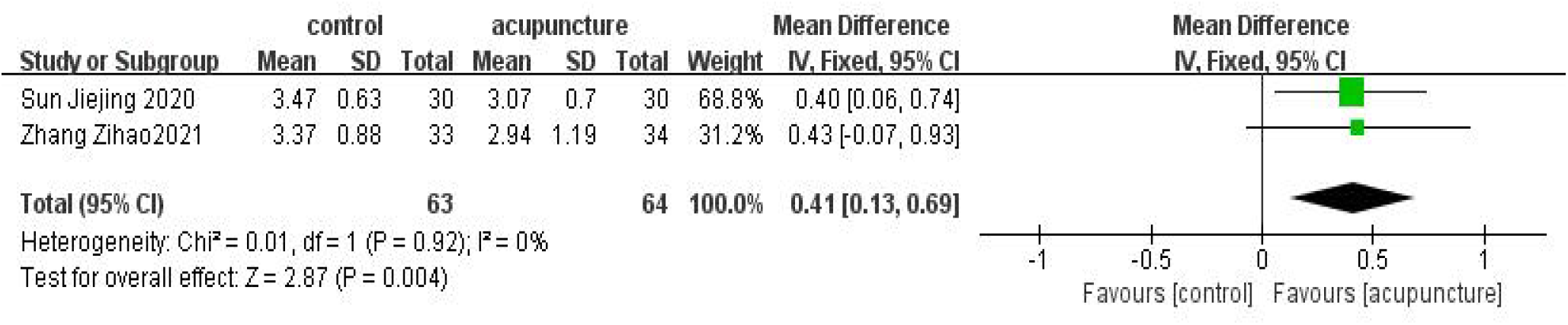
Forest plot of two groups of intervention 2-day pain levels

### 2.4 Publication bias

The total clinical efficacy of published bias analysis for all outcome indicators was 6 articles, with a nearly symmetrical distribution of funnel points, as shown in Figure 6.

**Figure 6.**
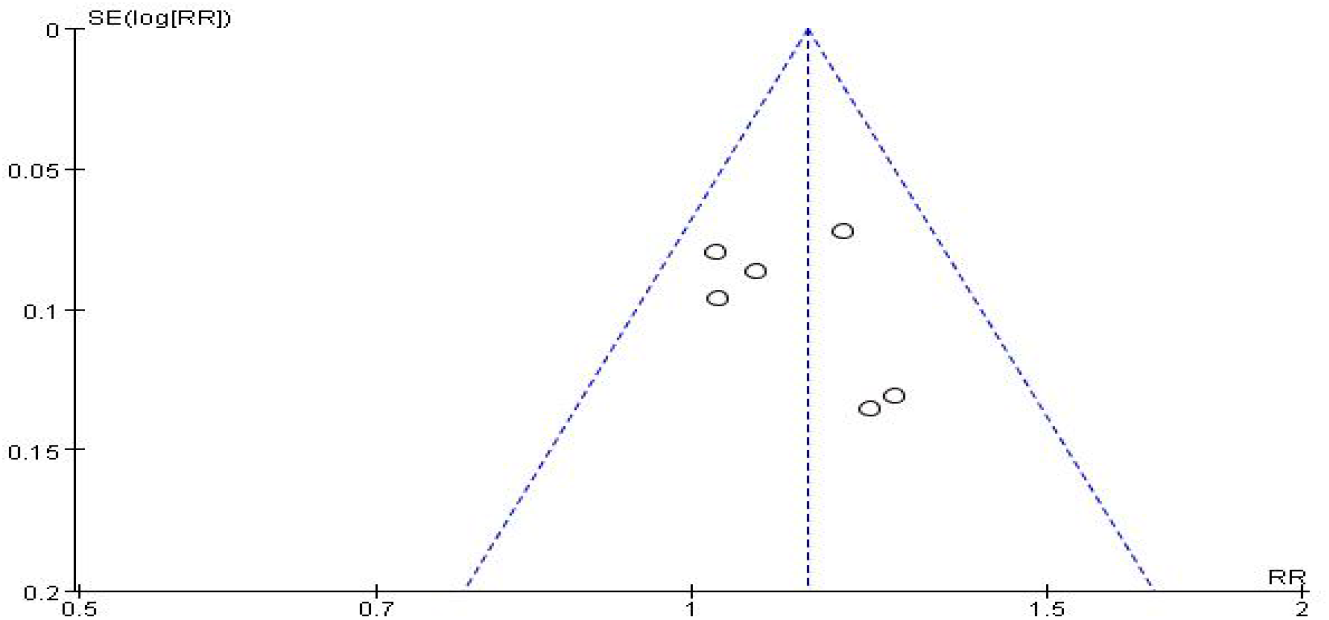
Funnel Diagram of Total Clinical Efficiency

### 2.5 Safety evaluation

Adverse reactions, including nausea, vomiting, dizziness and headaches, were reported in only two of the studies[12][18] included in the safety evaluation. The results showed that the observed group had fewer adverse reactions than the control group, and the difference was statistically significant (P < 0.05), meaning that Chinese or Western medicine caused many adverse reactions compared to acupuncture, as shown in Figure 7.

**Figure 7.**
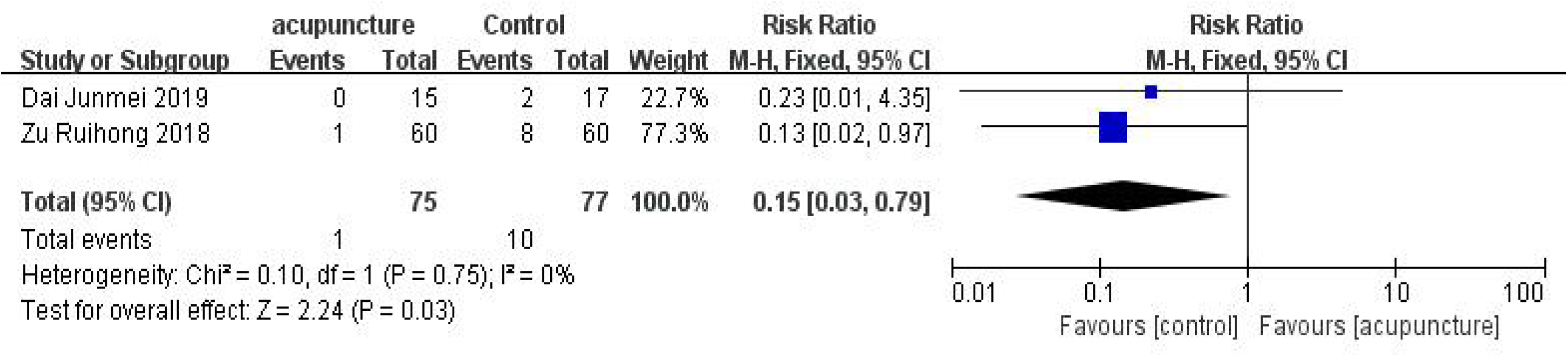
Forest plog of two sets of adverse reactions

## 3. discussion

Hemorrhoids have gradually become a common disease in today’s society, “ten people nine hemorrhoids’’ is no longer false rumors. The cause of hemorrhoids is closely related to daily habits. Sedentary, standing for a long time, fatigue, pregnancy[21] and so on make the body in a fixed position for a long time, thus affecting blood circulation, so that the pelvic blood flow is slow and abdominal organ congestion, causing excessive hemorrhoid vein filling, varicose, bulge, vein wall tension to decrease and cause hemorrhoids[22]. Second, eating habits are also an important reason, long-term drinkers or people who like spicy food, because alcohol and spicy substances can stimulate the digestive tract mucous membranes, resulting in vasodilation, colon dysfunction, anal intestinal diseases significantly increased morbidity. The main manifestations of hemorrhoids are: intermittent defecation followed by blood, difficulty defecating, pain, etc. It even increases the risk of depression[23] for hemorrhoids, which grow in private parts of the body. Many people with hemorrhoids are too ashamed to let their doctors see what they have and delay their condition. The longer it goes on, the more serious the condition becomes, and the fear of the disease accumulates in the back of their minds, becoming an unspoken secret and easily a burden on their hearts. Surgery is still the main treatment. But postoperative complications are unavoidable.

This study systematically discusses the complication of postoperative pain. Meta-analysis showed no statistically significant difference in 2-hour pain levels compared to intervention [MD=0.01, 95% CI (−0.23, 0.24), P=0.95]. After 2 days of intervention, pain levels [MD=-0.41, 95% CI (−0.69, -0.13), P=0.004], the total efficacy between the two groups was compared [RR=1.14, 95% CI (1.06, 1.24), P= 0.001], and the difference was statistically significant (P < 0.05). Surface-acupuncture [24][25] treatment of postoperative pain in hemorrhoids increased the total clinical efficacy. However, there was no significant difference between acupuncture and traditional Chinese medicine and Western medicine during the short intervention period, and the advantages of acupuncture for postoperative pain were gradually increased with time of treatment. Acupuncture is simple, safe and effective, and the adverse reaction of western medicine is obviously higher than acupuncture. Proper postoperative rehabilitation exercises[26] and TCM diet care[27] also help relieve pain and promote wound healing. However, the study has some limitations, such as some of the research design flaws, interventions are not detailed description; Safety was not evaluated in most of the included studies; Efficacy indicators were inconsistent. High-quality randomized controlled trial studies will be conducted in the future with scientific design, complete reporting, and standard implementation.

Combining the meta-analysis results, the bias characteristics mentioned above and the limitations of this paper, we conclude that acupuncture can relieve the pain and improve the quality of life of patients after hemorrhoid surgery and is safe. Therefore, acupuncture can be recommended to relieve the symptoms of postoperative pain after hemorrhoid surgery. However, randomized controlled trials with multiple centers and large samples will be needed in the future to provide more robust evidence.

## Data Availability

All data produced in the present study are available upon reasonable request to the authors.

## Author Approval

Xinyan Zou: Methodology, Formal analysis, and Writing – Original Draft; Qiaoqiao Liu: Methodology and Writing – Review & Editing; Longxia Gao: Writing – Review & Editing; Hanqing Zhao: Conceptualization, Resources, Writing – Review & Editing and Funding acquisition.

## References

[1] Sandler RS, Peery AF. Rethinking What We Know About Hemorrhoids. ClinGastroenterolHepatol. 2019 Jan;17(1):8–15.

[2] Gkegkes ID, Dalavouras N, Iavazzo C, Stamatiadis AP. Sweetening … the pain: The role of sugar in acutely prolapsed haemorrhoids. Clin Ter. 2021 Nov 22;172(6):520–522.

[3] Huang H, Gu Y, Ji L, Li Y, Xu S, Guo T, Xu M. A NEW MIXED SURGICAL TREATMENT FOR GRADES III AND IV HEMORRHOIDS: MODIFIED SELECTIVE HEMORRHOIDECTOMY COMBINED WITH COMPLETE ANAL EPITHELIAL RETENTION. Arq Bras Cir Dig. 2021 Oct 15;34(2):e1594.

[4] McKeown DG, Goldstein S. Hemorrhoid Banding. 2022 Jul 25. In: StatPearls[Internet]. Treasure Island (FL): StatPearls Publishing; 2022 Jan–.

[5] Alvarez-Downing MM, da Silva G. ‘Bumps down under:’ hemorrhoids, skin tags and all things perianal. Curr Opin Gastroenterol. 2022 Jan 1;38(1):61–66.

[6] Wang, TH., Kiu, KT., Yen, MH. et al. Comparison of the short-term outcomes of using DST and PPH staplers in the treatment of grade III and IV hemorrhoids. SciRep 10, 5189 (2020).

[7] Sammour T. Pain After Hemorrhoidectomy. Dis Colon Rectum. 2022 Aug 1;65(8):951–952.

[8] Yue B, Wang Y, Zhang C, Ding Y, Liu Z. Efficacy of Shaobei injection in the treatment of grade II-III hemorrhoids and the effect on fibulin protein expression: A study protocol of a randomized controlled trial. Medicine (Baltimore). 2021 Nov 19;100(46):e27706.

[9] Ye S, Zhou J, Guo X, Jiang X. Three Acupuncture Methods for Postoperative Pain in Mixed Hemorrhoids: A Systematic Review and Network Meta-Analysis. Comput Math Methods Med. 2022 Sep 26;2022:5627550.

[10] Lin S, Zang M. Effectiveness of Mayinglong Musk Hemorrhoid Ointment on Wound Healing and Complications after Internal Hemorrhoid Ligation and External Hemorrhoidectomy. Evid Based Complement Alternat Med. 2022 Jun 9;2022:5630487.

[11] GU Jao. Clinical Study on Wrist-ankle Acupuncture on Relieving Pain after Operation of Mixed Hemorrhoids[J]. CHINESE MEDICINE MODERN DISTANCE EDUCATION OF CHINA, 2020;18(4):82–84.

[12] ZU Ruihong. Clinical Observation on Acupoint Application Combined with Acupuncture and Moxibustion on Postoperative Analgesia after Hemorrhoids Operation[J]. CJGMCM, 2018, 33(22): 3371–3373.

[13] HE Ying-hua, ZHI Jian-wen, JIA Fei, YUAN Liang, YANG Yi, WANG Xiao-feng. Clinical Effect of Auricular Needle-embedding Therapy in The Treatment of Postoperative Pain of Mixed Hemorrhoid[J]. Journal of Basic Chinese Medicine, 2018 sep, 24(09): 1280-1282–1308.

[14] Sun Jiejing. Observation on the efficacy of deep puncture at Baliao acupoint in the treatment of pain after exfoliation and internal ligation of mixed hemorrhoids[D]. Nanjing University of Chinese Medicine, 2020.

[15] Kaili Li. The clinical effect observation of Anshen,Liqi and Zhitong Acupuncture Therapy for Pain after Hemorrhoids Sergery[D]. Yunnan University of Chinese Medicine, 2021.

[16] Zhang Zihao. Clinical Study on Postoperative Pain and Complications of Mixed Hemorrhoids[D]. HuNan University of Chimese Medicine, 2021.

[17] Lei Hongfeng, Yunan, Huawei. Clinical Observation on Treatment of Anal Edema after External Dissection & Internal Ligature for Mixed Hemorrhoids by Acupuncture plus Heat-sensitization Moxibustion[J]. Proceeding of Clinical Medicine, 2019, 28(04): 263-264–313

[18] Dai Junmei, Guo Minghao. Clinical Observation of Acupoint Application Combined with Moxibustion in th Treatment of Pain after External Dissection & Internal Ligature[J]. Modern Digestion & Intervention, 2019,(S2): 2479

[19] Lin Dongyin, Yuan Changyu, Shi Aiqiong, Zhang Yanqin. Observation on the Effect of Moxibustion Box Acupoint Formula on Incision Pain after Hemorrhoids Operation[J]. Nursing and Rehabilitation, 2018, 17(03): 83–85.

[20] Liu Meizhen(1), Duan Huanhuan(2). Comparison of the Effects of Moxibustion Therapeutic Apparatus and Tramadol Sustained-release Tablets in the Treatment of Anal Pain after Mixed Hemorrhoids[J]. Chin J of Clinical Rational Drug Use, 2017; 10(19):81–82

[21] Poskus T, Sabonyte-Balsaitiene Z, Jakubauskiene L, Jakubauskas M, Stundiene I, Barkauskaite G, Smigelskaite M, Jasiunas E, Ramasauskaite D, Strupas K, Drasutiene G. Preventing hemorrhoids during pregnancy: a multicenter, randomized clinical trial. BMC Pregnancy Childbirth. 2022 Apr 30;22(1):374.

[22] Li Junlai. Analysis of Clinical Efficacy and Related Factors of Surgical Treatment of Hemorrhoids[J]. Journal of Clinical Medical, 2017, 4(16): 3017.

[23] Xiao Meiling. Effect of Comprehensive Pain Relief Nursing Mode on Postoperative Pain and Anxiety in Patients with Hemorrhoids Surgery [J]. ModDiagnTreat, 2022, 33(11): 1727–1729.

[24] LIU Jiguang, ZHANG Xiaojian, ZHANG Songtao. Effect of Acupuncture for Postoperative Recovery of Patients with Hemorrhoids[J]. LiaoNing Traditional Chinese Medicine, 2022, 49(03): 193–195.

[25] Xie Hui, Li Hekun, Gu Ziwei, Zhang Nan, Zhang Liancheng. Research Progress of Acupuncture Treatment for Hemorrhoids[J]. Chin J Acu and Mox, 2021, 10(03): 111–113.

[26] Hu Xuelian. Effect of Anus Lifting Combined with Fine Nursing on Pain and Rehabilitation of Patients with Hemorrhoids after Operation[J]. Acta Medicinae Sinica, 2022, 35(02): 146–148.

[27] WEI Yanyang, M. Chong’ai, LUO Yanjin, LI Man, LUO Mingkui. Study on the Traditional Chinese Dietary Intervention on Postoperative Rehabilitation of Hemorrhoid[J]. ChineseandForeign Medical Research, 2020, 18(35): 160–162.

